# Epigenome-wide association study of seizures in childhood and adolescence

**DOI:** 10.1101/19005116

**Authors:** Doretta Caramaschi, Charlie Hatcher, Rosa H. Mulder, Janine F. Felix, Charlotte A. M. Cecil, Caroline L. Relton, Esther Walton

## Abstract

The occurrence of seizures in childhood is often associated with neurodevelopmental impairments and school underachievement. Common genetic variants associated with epilepsy have been identified and epigenetic mechanisms have also been suggested to play a role. In this study we analysed the association of genome-wide blood DNA methylation with the occurrence of seizures in ∼800 children from the Avon Longitudinal Study of Parents and Children, UK, at birth (cord blood), during childhood and adolescence (peripheral blood). We also analysed the association between the lifetime occurrence of any seizures before age 13 with blood DNA methylation levels. We sought replication of the findings in the Generation R Study and explored causality using Mendelian randomization, i.e. using genetic variants as proxies. The results showed five CpG sites which were associated cross-sectionally with seizures either in childhood or adolescence (1-5% absolute methylation difference at p_FDR_<0.05), although the evidence of replication in an independent study was weak. One of these sites was located in the *BDNF* gene, which is highly expressed in the brain, and showed high correspondence with brain methylation levels. The Mendelian randomization analyses suggested that seizures might be causal for changes in methylation rather than vice-versa. In addition, seizure-associated methylation changes could affect other outcomes such as growth, cognitive skills and educational attainment. In conclusion, we present a link between seizures and DNA methylation which suggests that DNA methylation changes might mediate some of the effects of seizures on growth and neurodevelopment.

## INTRODUCTION

Seizures are episodes of abnormal excessive or synchronous neuronal activity in the brain that affect 1-14% of children under six years of age, with the highest incidence in underdeveloped and rural areas ^1, 2^. The peak age for seizures to occur is at 18 months of age and children that experienced seizures are at risk of developing epilepsy. Seizures and epilepsy are associated with neurodevelopmental conditions, such as autism spectrum disorders ^3^, attention-deficit hyperactivity disorder and cognitive impairment ^4-7^. Moreover, epilepsy with or without intellectual impairment is associated with low academic achievement ^8^.

Genome-wide association studies have identified genetic variants associated with epilepsy ^9, 10^, some of which are located in candidate genes for epilepsy, for instance those coding for ion-channel subunits, and are supported by other research in humans and other animals. As it is likely that other factors may also underlie the disease, it has been suggested that epigenetic mechanisms such as DNA methylation are also involved in the onset of seizures ^11^. In line with this hypothesis, genetic markers for epilepsy were found to be enriched in histone modification markers, suggesting epigenetic regulation of gene transcription ^9^. The association between seizures and DNA methylation has been investigated in studies involving humans and other animals, although these studies relied on small sample sizes or on a candidate gene approach. A recent study comparing blood DNA methylation in 30 adult patients with mesial temporal lobe epilepsy and 30 controls identified 216 differentially methylated sites between the two groups, including sites on genes involved in ion binding and metabolic activity ^12^. DNA methylation differences have also been observed in lymphoblastoid cell lines derived from epileptic patients globally and in the *BRD2* gene promoter ^13^. Another study that reanalysed these data discovered differential DNA methylation in non-coding RNAs ^14^. Furthermore, alterations in DNA methylation were present in the hippocampus of epileptic patients compared to controls ^15^. A study adopting a rat model of chronic epilepsy corroborated these findings by revealing genome-wide differences in DNA methylation compared to control rats ^16^.

Typically, in association studies it is difficult to assess the causality of any identified association due to the potential for confounding and/or for reverse causation. Socioeconomic status, for instance, is associated with genome-wide changes in DNA methylation ^17^ and with increased risk for seizures/epilepsy ^18^, suggesting that socioeconomic factors could be confounding the association between DNA methylation and seizures. With respect to reverse causation, case-control studies that examined DNA methylation after epilepsy had already been diagnosed might have observed changes that were directly caused by the seizure events. For instance, laboratory animal studies have shown altered gene expression after induced seizures ^19^. Mendelian randomization, a technique that uses the genetic information associated with an exposure to estimate the causal effect of the exposure on an outcome, can circumvent these limitations under certain assumptions ^20^.

In this study, we (1) investigated the genome-wide association of DNA methylation with the occurrence of seizures from birth throughout childhood and adolescence in peripheral blood samples from a prospective birth cohort; (2) carried out replication analyses in an independent study sample; (3) explored the correspondence with brain tissue DNA methylation at the same genomic locations to investigate whether these associations have neurodevelopmental relevance; and (4) performed bi-directional Mendelian randomization to unravel causal links between peripheral blood DNA methylation and seizures. Finally, we explored the potential health consequences of a seizure-associated DNA methylation profile. For an overview on our analysis plan, see Figure 1.

**Figure 1.**
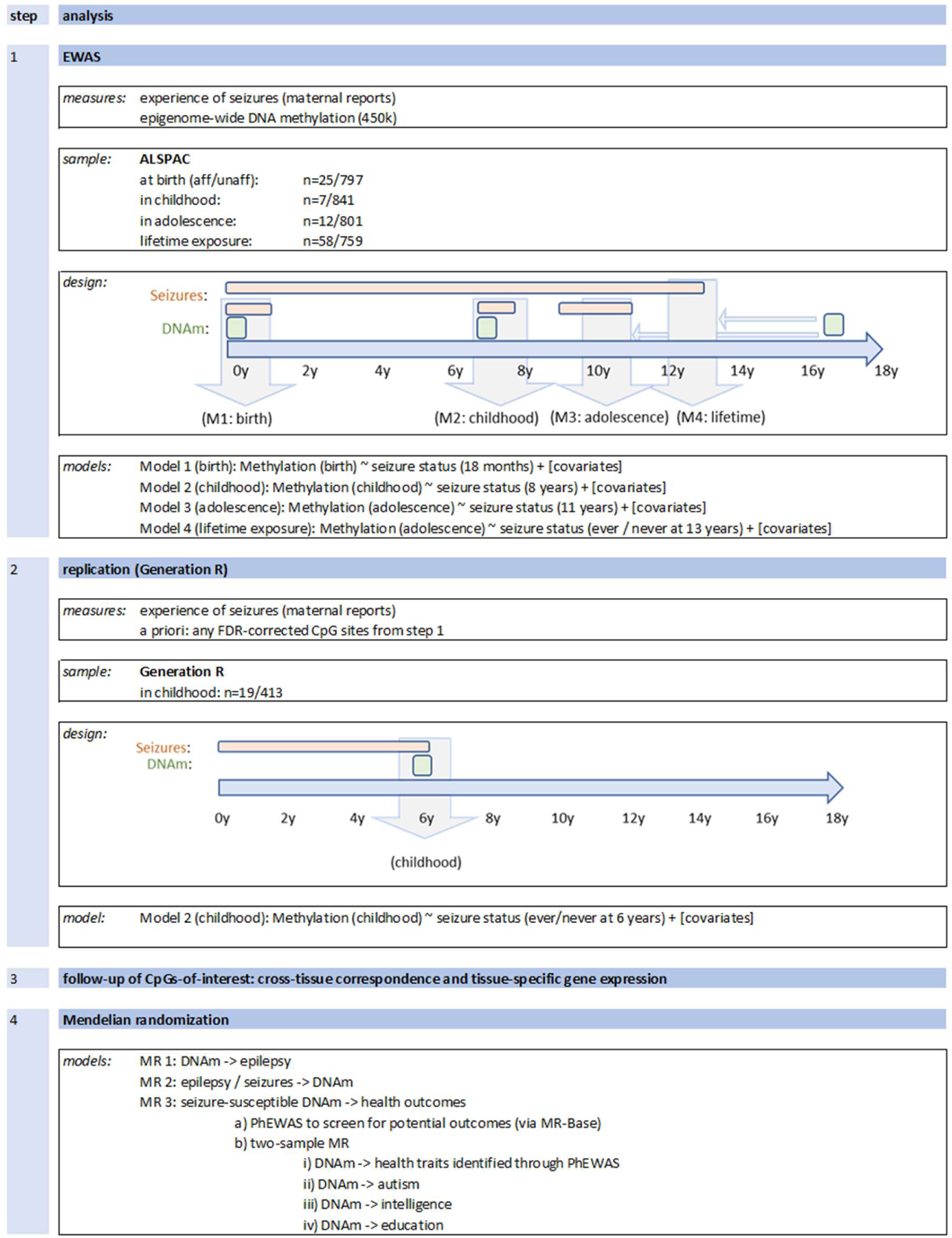
Analysis overview.

## MATERIALS AND METHODS

### Study Population

The discovery analyses were conducted in the Avon Longitudinal Study of Parents and Children (ALSPAC), a large prospective cohort study that recruited 14,541 pregnant women, resident in Avon, UK with expected delivery dates between the 1^st^ of April 1991 and the 31^st^ of December 1992 ^21, 22^. Of these initial pregnancies, there were 14,062 live births and 13,988 children who were alive at 1 year of age. The study website contains details of all the data that are available through a fully searchable data dictionary (http://www.bris.ac.uk/alspac/researchers/data-access/data-dictionary/).

Written informed consent has been obtained for all ALSPAC participants. Ethical approval for the study was obtained from the ALSPAC Ethics and Law Committee and the Local Research Ethics Committees.

### Seizure Data

Questionnaire data were taken from ALSPAC questionnaires administered to the mothers at four time points: 18 months, 8 years, 11 years and 13 years. The following questions were asked at each of these ages:

<u>18 months</u>: ‘Has he/she ever had any form of convulsion/fit/seizure or other turn in which consciousness was lost or any other part of the body made an abnormal movement?’,

<u>8 years</u>: ‘Has child had a convulsion/fit/seizure since 7^th^ birthday?’,

<u>11 years</u>: ‘Has child had a convulsion/fit/seizure where consciousness or abnormal movement was lost since 9^th^ birthday?’, and

<u>13 years</u>: ‘Has she ever had a seizure, fit or a convulsion?’.

This allowed us to address proximal and distal (i.e. lifetime until age 13) associations between DNA methylation and experience of seizures.

### DNA Methylation Data

In ALSPAC, blood from 1018 mother–child pairs were selected for analysis as part of the Accessible Resource for Integrative Epigenomic Studies (ARIES, http://www.ariesepigenomics.org.uk/) ^23^. Following DNA extraction, samples were bisulphite converted using the Zymo EZ DNA Methylation™ kit (Zymo, Irvine, CA, USA) and genome-wide methylation was measured using the Illumina Infinium HumanMethylation450 (HM450) BeadChip. The arrays were scanned using an Illumina iScan, with initial quality review using GenomeStudio. ARIES was pre-processed and normalised using the *meffil* R package ^24^. ARIES consists of from mother-child pairs measured at five time points (three time points for children: birth, childhood and adolescence; and two for mothers: during pregnancy and at middle age), although only children’s profiles were used in the current study. Low quality profiles were removed from further processing, and the remaining 4593 profiles were normalised using the Functional Normalization algorithm ^25^ with the top 10 control probe principal components. Full details of the pre-processing and normalization of ARIES has been described previously ^24^. Further pre-processing specific to the current study included removal of probes not passing background detection (p>0.05) and probes on the X or Y chromosome. To reduce the impact of outliers, we set methylation data points outside the 3 x inter-quartile-range to missing. The total number of probes available for analyses were N=468,828 at birth; N= 471,092 at childhood and N=470,480 at adolescence.

### Epigenome-wide association analyses

In ALSPAC, the final sample size at birth was N=822 (25 cases and 797 controls); at childhood N=848 (7 cases and 841 controls) and N=813 (12 cases and 801 controls) at adolescence. Only 2-3 cases overlapped across time points (Supplemental Material SM Figure 1). Fifty-eight out of a total of N=817 adolescents reported a lifetime experience of seizures.

We carried out four analyses. In analysis 1-3, we modelled methylation at birth (Model 1), childhood (Model 2) and adolescence (Model 3) as the outcome and seizure status (measured closest to each methylation time point) as the exposure. To investigate the temporal sensitivity of associations, we ran a final analysis (Model 4), in which we modelled lifetime seizure status (ever / never) at 13 years as the exposure and methylation at adolescence as the outcome (Model 4). In all models (including birth), methylation was defined as the outcome regardless of temporal order to keep model estimates consistent and comparable. These EWAS were carried out in R version 3.3.1 using the CpGassoc package^26^.

All models were adjusted for age (when DNA methylation samples were taken), sex, prenatal maternal smoking (Yes/No) and maternal education (university degree Yes/No), each derived from ALSPAC maternal and childhood questionnaires. The model using cord blood data was additionally adjusted for gestational age and birthweight. Unknown confounders and batch were adjusted using surrogate variable analysis ^27^. Additionally, we adjusted for cell counts using the Houseman method for the childhood and adolescence timepoints^28^ and the Andrews and Bakulski method for cord blood^29^.

To summarize, the following models were applied:

Model 1: Methylation (cord) ∼ seizure status (18 months) + age + sex + birthweight + gestational age + maternal prenatal smoking + maternal education + nucleated red blood cells + granulocytes + monocytes + natural killer cells + B cells + CD4(+)T cells + CD8(+)T cells + SV1 + … + SV15

Model 2: Methylation (childhood) ∼ seizure status (8 years) + age + sex + maternal prenatal smoking + maternal education + granulocytes + monocytes + natural killer cells + B cells + CD4(+)T cells + CD8(+)T cells + SV1 + … + SV13

Model 3: Methylation (adolescence) ∼ seizure status (11 years) + age + sex + maternal prenatal smoking + maternal education + granulocytes + monocytes + natural killer cells + B cells + CD4(+)T cells + CD8(+)T cells + SV1 + … + SV14

Model 4: Methylation (adolescence) ∼ seizure status (ever / never at 13 years) + age + sex + maternal prenatal smoking + maternal education + granulocytes + monocytes + natural killer cells + B cells + CD4(+)T cells + CD8(+)T cells + SV1 + … + SV14

To correct to multiple testing, we present both Bonferroni and FDR-corrected results.

### Replication analyses

All CpG sites that were associated with seizures below at least an FDR-correction threshold in ALSPAC were analysed in an independent cohort to assess replication. The Generation R Study is a population-based prospective cohort study conducted in Rotterdam, the Netherlands, that recruited 9778 pregnant women with an expected delivery date between April 2002 and January 2006. A total of 9749 children were born from these pregnancies and extensive data and biological samples are available from the children and their mothers ^30^. DNA methylation was measured in peripheral blood of 469 children aged 6 years, using the Infinium HumanMethylation450 (HM450) BeadChip as in ALSPAC. Preparation and normalization of the BeadChip array was performed according to the CPACOR workflow in R ^31^ and methylation data points lower than the 25th percentile – 3 x IQR and higher than the 75th percentile + 3 x IQR were excluded. In Generation R, seizure events were measured using the answer “yes” to the question “During the past 5/6 years, did your child ever have a seizure/febrile convulsion?” asked to the mothers when the children were 6 years of age. The final sample size was N=432, with 19 participants affected by seizures and 413 unaffected (SM Table 1). Linear models similar to Model 2 were run in Generation R on EWAS FDR-corrected methylation sites and on *BDNF* probes. The covariates were measured and categorised similarly to the analyses carried out in ALSPAC. The results in ALSPAC and Generation R where also meta-analysed using METAL ^32^, using inverse variance weighting.

**Table 1.** ARIES sample characteristics.

| Characteristic | Birth |  |  | Childhood |  |  | Adolescence |  |  | Adolescence (Lifetime exposure) |  |  |
| --- | --- | --- | --- | --- | --- | --- | --- | --- | --- | --- | --- | --- |
|  | with seizures | without seizures | total | with seizures | without seizures | total | with seizures | without seizures | total | with seizures (ever) | without seizures (ever) | total |
| N | 25 | 797 | 822 | 7 | 841 | 848 | 12 | 801 | 813 | 58 | 759 | 817 |
| Sex (%F) | 44% | 51% | 51% | ND | 50% | 51% | ND | 51% | 52% | 52% | 51% | 51% |
| Age at methylation in years (SD) | NA |  |  | 7.69 (0.36) | 7.44 (0.13) | 7.45 (0.14) | 17.65 (0.86) | 17.11 (1.04) | 17.12 (1.04) | 17.16 (1.01) | 17.10 (1.05) | 17.10 (1.04) |
| Age at seizure assessment in years (SD) | 1.54 (0.12) | 1.52 (0.06) | 1.52 (0.07) | 8.60 (0.03) | 8.66 (0.16) | 8.66 (0.16) | 11.70 (0.08) | 11.71 (0.12) | 11.71 (0.12) | 13.12 (0.09) | 13.15 (0.17) | 13.43 (1.05) |
| Birthweight in g (SD) | 3498.80 (545.54) | 3495.08 (478.18) | 3495.20 (480.00) | NA |  |  | NA |  |  | NA |  |  |
| Gestational age in weeks (SD) | 39.80 (1.50) | 39.55 (1.48) | 39.56 (1.48) |  |  |  |  |  |  |  |  |  |
| Maternal smoking (%Y) | ND | 13% | 14% | ND | 12% | 13% | ND | 13% | 13% | 14% | 13% | 13% |
| Maternal education (%Uni) | 32% | 20% | 21% | ND | 21% | 21% | ND | 22% | 22% | 28% | 22% | 22% |

ND=Not disclosed due to cohort restrictions. NA= Not applicable.

### Mendelian randomization analyses

To assess the causal link between DNA methylation and the occurrence of seizures we performed two-sample Mendelian randomization (MR) using the EWAS results for the CpG sites with FDR-corrected p-values<0.05 in ALSPAC. Two-sample MR was performed using the MR-Base online platform (http://www.mrbase.org/, last accessed 06-06-2018) ^33^, the MRInstruments R package (https://github.com/MRCIEU/MRInstruments, last accessed 06-06-2018) and the TwoSampleMR R package (https://github.com/MRCIEU/TwoSampleMR, last accessed 06-06-2018).

Three MR analyses were performed. (1) To investigate the causal effect of DNA methylation on the risk of epilepsy we performed two-sample MR with DNA methylation as exposure and a diagnosis of epilepsy as outcome. For the genotype-exposure associations we searched for methylation quantitative trait loci (mQTLs), i.e. genetic variants that are associated in *cis* with DNA methylation (i.e. within 1Mb either side from the CpG site), using the mQTL database (http://www.mqtldb.org/, last accessed 06-06-2018)^34^, restricting the search to the time point, at which the CpG site was associated with seizure status. For the genotype-epilepsy association we used the summary statistics for ICD-9 and ICD-10 codes for epilepsy or seizure in MR-Base generated on UKBiobank data (last accessed 04-12-2018). (2) To analyse the causal effect of risk for seizure/epilepsy on DNA methylation (i.e. reverse causation) we performed two-sample MR with epilepsy diagnosis or febrile/vaccine-related seizures as exposure and DNA methylation as outcome. For the genotype-exposure associations we used the summary statistics for the genome-wide significant SNPs from a published GWAS meta-analysis on all epilepsies, focal epilepsy and genetic generalized epilepsy ^10^ and from a published GWAS on febrile and MMR-vaccine-related seizures (focussing on 6 replicated genome-wide significant

SNPs in Table1 of the original article) ^35^. Odds ratios (OR) and confidence intervals were reverted to log-odds and standard errors to use in the two-sample MR analysis. +The summary statistics for the genotype-outcome associations were drawn from the mQTL database (http://www.mqtldb.org/, last accessed 06-06-2018)^34^. (3) To analyse the causal effect of seizure-susceptible DNA methylation (exposure) on other health outcomes we first performed a hypothesis-free PheWAS using the mQTLs for CpG sites associated with seizures and the MR-Base online PheWAS tool to screen for potentially affected health outcomes. Then, we performed two-sample MR with methylation as exposure and the health outcomes identified by the PheWAS to estimate the magnitude of the effect. We also performed a hypothesis-driven two-sample MR on other neurodevelopmental outcomes previously found to associate with seizures, i.e. autism, intelligence and education.

## RESULTS

For a flowchart and overview of all results, see SM Figure 2.

**Figure 2.**
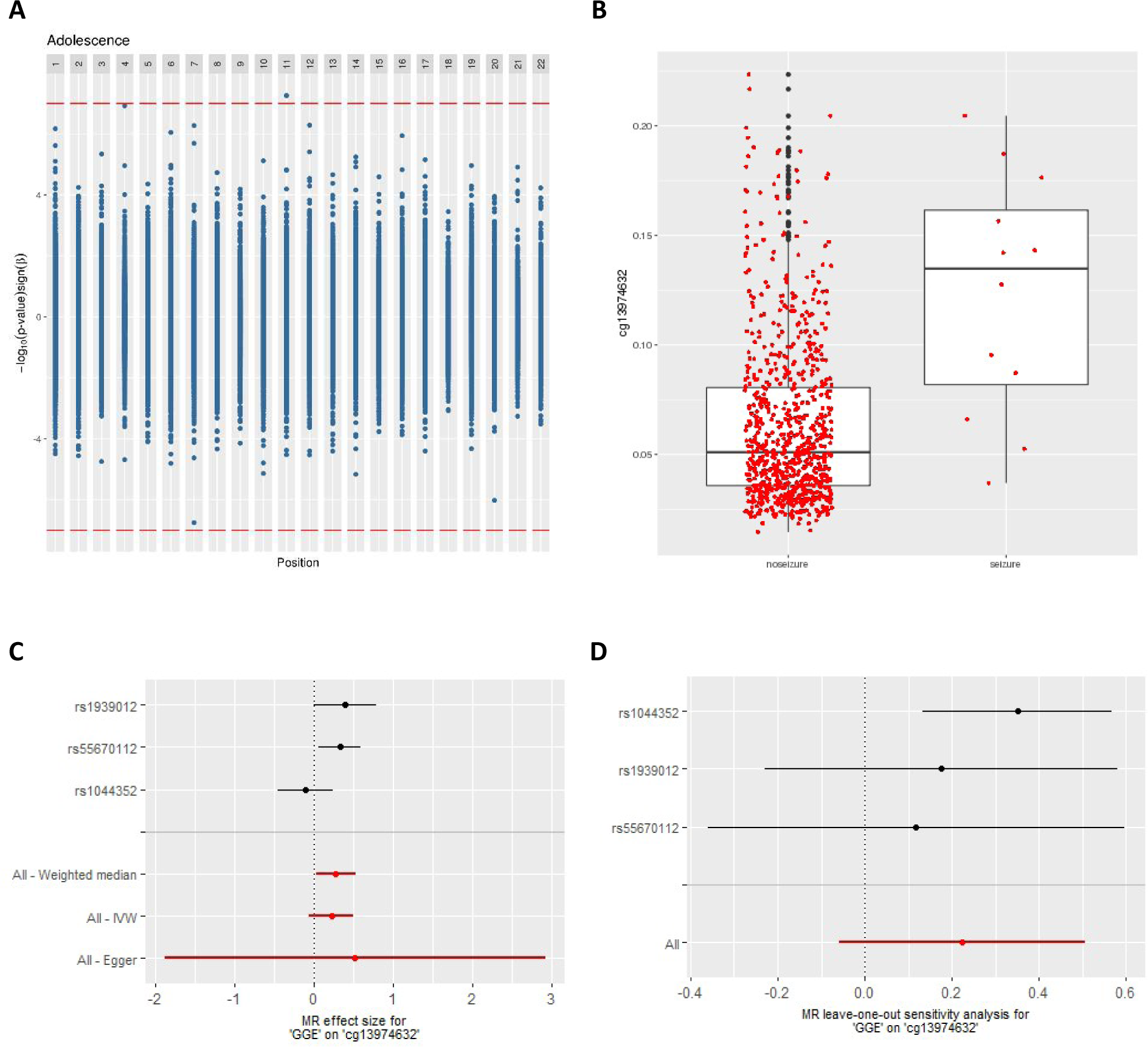
A) Miami plot displaying the EWAS results by chromosome in adolescence. Positive values on y-axis indicate −log(P-values) of hypermethylated sites, whereas negative values on the y-axis indicate −log(P-values) of hypomethylated sites (the sign of the y-axis values have been changed to reflect this). Bonferroni cut-off line in red. B) Boxplot of methylation levels at the *BDNF*-linked CpG cg13974632 (adjusted for covariates). C) Causal estimate for the effect of genetic generalized epilepsy on cg13974632 (*BDNF*). Individual SNP results in black and overall causal estimates in red. D) Leave-one-out analysis of the causal estimate for the effect of genetic generalized epilepsy on cg13974632 (*BDNF*). IVW=Inverse variance weighted.

### Sample description

At 18 months, there were n=25 children with the experience of seizures since birth and n=797 without (Table 1). Groups were comparable with respect to sex, birthweight, gestational age as well as maternal education and smoking behaviour during pregnancy. During childhood, n=7 children had experienced seizures between 7 and 8 years of age while 841 had not. Age at blood draw was slightly higher in the children with seizures (Table 1). Approaching adolescence, seizures had been reported in n=12 children, while n=801 children had not experienced seizures between 9 and 11 years of age. There were slightly more females in the seizure group. Fifty-eight adolescents reported a lifetime experience of seizures, while n=759 had never experienced seizures. Groups were comparable with respect to maternal smoking behaviour, maternal education and age at blood draw. For correlation plots on seizure rates and all covariates included in the final models specific to each time point, see SM Figure 3.

### Epigenome-wide association analyses

We did not identify any CpG sites that fell below a Bonferroni or FDR adjusted P-value threshold at birth (Table 2 and SM Figure 4). In childhood, two CpG sites associated with seizure status after FDR correction (cg10541930: beta = −0.010, SE = 0.002, uncorrected P-value = 4.32 × 10^−8^ and FDR = 0.020; cg25557432: beta = 0.014, SE = 0.003, uncorrected P-value = 1.82 × 10^−7^ and FDR = 0.043). The first CpG is located in an intergenic region, at the transcription start site for a non-coding RNA, while the second lies upstream of the gene *MACROD2* involved in DNA repair.

**Table 2.** EWAS results. Top CpG sites for the three time-points: birth, childhood and adolescence (cross-sectional and lifetime exposure).

| Time point | probeID | Beta | SE | P-value | FDR | Bonferroni | Chromosome | Position | Gene |
| --- | --- | --- | --- | --- | --- | --- | --- | --- | --- |
| Birth | cg07504545 | -0.042 | 0.008 | 2.32E-07 | 0.109 | 0.109 | 1 | 203456019 | *PRELP* |
|  | cg04977770 | -0.054 | 0.011 | 7.15E-07 | 0.168 | 0.335 | 17 | 79846763 | *THOC4* |
|  | cg23625106 | -0.023 | 0.005 | 1.47E-06 | 0.193 | 0.689 | 8 | 61789727 |  |
|  | cg08461451 | -0.036 | 0.008 | 1.73E-06 | 0.193 | 0.812 | 19 | 2295092 | *LINGO3* |
|  | cg06367149 | 0.057 | 0.012 | 2.08E-06 | 0.193 | 0.973 | 15 | 61254575 | *RORA* |
|  | cg13396019 | -0.026 | 0.005 | 2.57E-06 | 0.193 | 1.000 | 1 | 220564510 |  |
| Childhood | cg10541930 | -0.010 | 0.002 | 4.32E-08 | 0.020 | 0.020 | 10 | 131909085 |  |
|  | cg25557432 | 0.014 | 0.003 | 1.82E-07 | 0.043 | 0.086 | 20 | 13976117 | *MACROD2* |
|  | cg11317171 | 0.002 | 0.000 | 1.34E-06 | 0.190 | 0.633 | 19 | 5623025 | *SAFB2;SAFB* |
|  | cg06447795 | 0.090 | 0.019 | 1.62E-06 | 0.190 | 0.761 | 20 | 36793996 | *TGM2* |
|  | cg02389501 | 0.078 | 0.016 | 2.59E-06 | 0.191 | 1.000 | 7 | 93757948 |  |
|  | cg13647052 | 0.018 | 0.004 | 2.82E-06 | 0.191 | 1.000 | 12 | 2800382 | *CACNA1C* |
| Adolescence | cg13974632 | 0.053 | 0.010 | 5.55E-08 | 0.026 | 0.026 | 11 | 27740813 | *BDNF* |
|  | cg15810326 | 0.014 | 0.003 | 1.19E-07 | 0.028 | 0.056 | 4 | 148605127 | *PRMT10* |
|  | cg16983916 | -0.056 | 0.011 | 1.79E-07 | 0.028 | 0.084 | 7 | 156159713 |  |
|  | cg11510269 | 0.007 | 0.001 | 5.18E-07 | 0.050 | 0.244 | 12 | 123011761 | *RSRC2;KNTC1* |
|  | cg17130518 | 0.052 | 0.010 | 5.33E-07 | 0.050 | 0.251 | 7 | 76825482 | *CCDC146;FGL2* |
|  | cg06222062 | 0.095 | 0.019 | 6.79E-07 | 0.053 | 0.319 | 1 | 20396690 | *PLA2G5* |
| Lifetime | cg20162381 | 0.004 | 0.001 | 9.22E-07 | 0.159 | 0.434 | 12 | 3310097 | *TSPAN9* |
|  | cg20156774 | -0.028 | 0.006 | 1.52E-06 | 0.159 | 0.717 | 6 | 32803500 | *TAP2* |
|  | cg12888660 | 0.003 | 0.001 | 1.72E-06 | 0.159 | 0.811 | 12 | 122018449 | *KDM2B* |
|  | cg15031661 | 0.006 | 0.001 | 1.91E-06 | 0.159 | 0.897 | 1 | 240256603 | *FMN2* |
|  | cg17233506 | -0.031 | 0.006 | 2.21E-06 | 0.159 | 1.000 | 17 | 46608293 | *HOXB1* |
|  | cg18108087 | 0.010 | 0.002 | 2.32E-06 | 0.159 | 1.000 | 1 | 59762112 | *FGGY* |

In adolescence, we found one CpG site that fell below a Bonferroni-adjusted P-value < 0.05: cg13974632 (beta = 0.053, SE = 0.010, uncorrected P-value = 5.55 × 10^−8^ and FDR = 0.026), while another two CpG sites passed an FDR threshold only (cg15810326: beta = 0.014, SE = 0.003, uncorrected P-value = 1.19 × 10^−7^ and FDR = 0.028; cg16983916: beta = 0.056, SE = 0.011, uncorrected P-value = 1.79 × 10^−7^ and FDR = 0.028; Figure 2A). The first CpG site is located in the first exon of the brain-derived neurotrophic factor (*BDNF)*. The experience of seizures was associated with increased DNA methylation at this site (Figure 2B). The other two CpG sites were located in the first exon of Protein Arginine Methyltransferase 10 (*PRMT10*) and in an intergenic region, respectively.

No CpG site could be identified to be associated with lifetime seizure experience. Inspection of QQ plots and lambdas close to 1 gave little indication for an inflation of test statistics (SM Figure 5).

### Replication in the Generation R Study

For replication, we focused on five CpG sites: two that passed FDR correction in childhood and three that passed FDR correction in adolescence. As one of the CpG sites in adolescence was located in the gene *BNDF*, which is a key driver in neuronal growth and has been repeatedly linked to epilepsy ^36, 37^, we expanded our search space to include all CpG sites annotated to *BDNF* (n=73).

None of the five CpG sites were associated with seizures in Generation R during childhood. Although the direction of effect suggested some degree of concordance, all but one beta coefficient were a factor of 10 smaller and P-values ranged between 0.1 and 0.8. (SM Table 2 and SM Figure 6A). When ALSPAC and Generation R results for these five probes where meta-analysed together, all CpGs except for cg16983916 showed evidence of methylation differences (p-value<0.05/77=0.0006, SM Table 2).

Investigating all 73 CpG sites annotated to *BDNF*, none replicated based on a correction for 73 tests. We observed only a weak correlation of all 73 regression betas between cohorts (rho=0.046, p-value =0.70 based on adolescence results in ALSPAC and childhood results in Generation R; SM Table 2 and SM Figure 6B). However, five CpG sites were significant at a nominal level. Furthermore, considering that the whole *BDNF* region spans almost 68k base pairs, we did note that one CpG site (cg25412831), which passed a nominal significance level (although in opposite direction) in Generation R, was only 1,325 base pairs away from the main *BDNF*-associated probe in ALSPAC (cg13974632). When ALSPAC and Generation R results for these 73 probes where meta-analysed together, there was evidence for methylation differences in 2 CpGs (p<0.05/77=0.0006, SM Table 2). These where cg13974632, the top hit from the EWAS, and cg15313332, 20Kb upstream.

### Cross-tissue concordance in DNA methylation

We queried three independent databases to investigate blood-brain concordance in DNA methylation for all five CpG sites that passed FDR correction in ALSPAC. Based on data from more than 122 pre-mortem blood samples and paired post-mortem brain tissue ^38^, cross-tissue correlation was strongest for BDNF_cg13974632_ (r=0.39 between blood and brain tissue from the entorhinal cortex, followed by tissue from the prefrontal cortex (r=0.27); SM Figure 7A-E. Compared to BDNF_cg13974632_, the remaining four CpG sites showed correlations, which were generally weaker for tissue from the prefrontal and entorhinal cortex. Cross-tissue correlations based on a smaller sample of 16 individuals ^39^ reported different correlation profiles (SM Figure 7F-G).

Cross-tissue blood-brain correlations in a dataset of 12 epilepsy patients ^40^ were consistent with the larger dataset, although not discriminating between brain regions (BDNF_cg13974632:_ rho=0.28, p=0.42; PRMT10_cg15810326_: rho-0.0, p=0.94; cg16983916: rho=0.09, p=0.81; cg10541930: rho=0.32, p=0.36; MACROD2_cg25557432_: rho=0.45, p=0.19).

Based on data available via the Genotype-Tissue Expression (GTEx; www.gtexportal.org) project, we investigated tissue-specific gene expression for genes linked to the five FDR-corrected CpG sites. *BDNF* appeared to be expressed in brain and other tissues with highest expression in the cerebellum, while very low expression was found in blood. *MACROD2* is predominately expressed in lymphocytes; *PRMT10* is mainly expressed in the ovaries (SM Figure 8).

### Mendelian randomization analyses

Uni- and bi-directional two-sample MR was performed to investigate the effect of DNA methylation on seizure occurrence and the effect of seizures on DNA methylation. We used genetic associations with febrile and vaccine-related seizures and genetic associations with epilepsy. The latter more generally includes the types of seizures observed in our study (e.g. febrile and non-febrile) ^41^. As full genome-wide summary statistics were only available for epilepsy, but not for febrile and vaccine-related seizures, we could not perform the two-sample MR to estimate the causal effects of methylation on seizures. We identified only one *cis*-mQTL, which could be used as instrument for DNA methylation. In detail, the SNP rs10258194 was associated in *cis* with cg16983916 (effect allele=T, beta=0.25, SD=0.04, p=2.33 × 10^−10^) after excluding other SNPs due to linkage disequilibrium. For the other CpG sites, either *trans*-associations (further than 1 Mb from the CpG site) or no associations were identified. Two-sample MR indicated only weak evidence for causal effects of DNA methylation at cg1698369 (SM Table 3) on epilepsy.

**Table 3.** Mendelian randomization analysis. Effects of DNA methylation at cg16983916 on non-epilepsy outcomes (PheWAS and candidate outcomes).

| Outcome | Reference | N | Beta<sup>a</sup> | S.E. | P-value (uncorrected) |
| --- | --- | --- | --- | --- | --- |
| Weight | UK Biobank <sup>b</sup> | 336227 | 0.016 | 0.009 | 0.067 |
| Trunk fat-free mass | UK Biobank <sup>b</sup> | 331030 | 0.025 | 0.006 | 3.73 x 10<sup>-05</sup> |
| Standing height | UK Biobank <sup>b</sup> | 336474 | 0.029 | 0.007 | 2.90 x 10<sup>-05</sup> |
| Years of schooling | Okbay et al., 2016 <sup>49</sup> | 182286 | 0.008 | 0.008 | 0.317 |
| Childhood intelligence | Benyamin et al., 2014 <sup>50</sup> | 12441 | 0.013 | 0.051 | 0.797 |
| Autism | Smoller et al., 2013 <sup>51</sup> | 29415 | -0.008 | 0.113 | 0.946 |
| Fluid intelligence score | UK Biobank <sup>b</sup> | 108818 | 0.119 | 0.036 | 0.001 |
| College or University degree | UK Biobank <sup>b</sup> | 334070 | 0.013 | 0.004 | 0.004 |

<sup>a</sup> Wald ratio
<sup>b</sup> Unpublished summary statistics from the UK Biobank. https://github.com/Nealelab/UK_Biobank_GWAS and http://www.nealelab.is/blog/2017/9/11/details-and-considerations-of-the-uk-biobank-gwas

For the reverse (i.e. epilepsy/seizure affecting DNA methylation), there were 9 SNPs to be used as instruments for epilepsy from a previous GWAS meta-analysis on all epilepsies, focal epilepsy and genetic generalized epilepsy (GGE), although only up to four SNPs were used in any one analysis due to availability of summary statistics. Six SNPs were identified as instruments for febrile/vaccine-related seizures, although only 5 were used. SM Table 4 shows the results of the two-sample MR analysis performed using different methods to investigate the causal effect of epilepsy on DNA methylation at the five CpG sites identified in the EWAS. For cg13974632 (*BDNF*) there was some evidence for a positive association of GGE with increased DNA methylation using the weighted median method (Figure 2 C-D). All methods, including MR-Egger, suggested a positive effect of GGE on cg13974632, although these analyses were based on only 3 genetic instruments and confidence intervals were large, particularly for MR-Egger. The leave-one-out analysis indicated that this effect was not driven by a particular genetic variant, providing little evidence for a violation of MR assumptions. This association did not survive a correction for multiple testing and seemed to be specific for GGE (i.e., the effect was not reproduced using focal epilepsy or “any epilepsy” as an exposure). There was no evidence for associations with any other CpG sites.

The two-sample MR analysis of the effects of febrile and vaccine-related seizures on methylation did not show enough evidence of a causal association (SM Table 5).

To test the effect of seizure-associated methylation on other health outcomes we first scanned for potentially relevant health traits by performing a PheWAS (association of genotype with all available outcomes) using the only mQTL available, rs10258194. As we used only one instrument, we could not differentiate whether the associations were due to causal effects or horizontal pleiotropy (i.e. the genetic variant has an effect on the health outcome outside of its effect on DNA methylation at the specific CpG). The analysis revealed strong evidence (Bonferroni-corrected p<0.05/776) for associations between rs10258194 and birth weight, standing height and trunk fat-free mass (SM Table 6). The two-sample MR analysis (Table 3) confirmed a positive, potentially causal, association of seizure-related DNA methylation at cg16983916 with trunk fat-free mass and standing height. Since a causal effect of seizures on birth weight is less plausible due to the temporal discordance of seizures occurring after birth, we performed two-sample MR on adult weight instead of birth weight (although in the screening PheWAS, adult weight was not associated with rs10258194 after multiple comparison correction) and found weak evidence of a positive association. There was some evidence of a positive association of methylation with fluid intelligence score and college or university qualification based on data from UK Biobank. However, we could not replicate these results based on data from other studies (Table 3). There was no strong evidence for an association with autism.

## DISCUSSION

In this study, we observed associations between blood DNA methylation and the occurrence of seizures in a longitudinal pregnancy cohort study based in the UK. Effects were specific to childhood and adolescence, with little evidence for a relationship at birth or for lifetime exposure to seizures. During adolescence, one of the strongest findings was linked to the *BDNF* gene. However, associations did not replicate in an independent study sample based in the Netherlands. Follow-up analyses showed correspondence between blood and brain DNA methylation levels in the *BDNF* gene and high expression across several brain regions. Mendelian randomization analyses suggest a potential causal effect of seizures on DNA methylation in the *BDNF* gene and potential negative consequences for growth and neurodevelopment via DNA methylation in an intergenic region on chromosome 7. Although the implications of the association with *BDNF* are interesting, the association was not replicated. The results are summarized in SM Figure 2.

This study has a number of strengths. First, in both ALSPAC and Generation R cohorts the information on seizures was provided by the parents near the time of occurrence, therefore reducing measurement error and the possibility of recall bias. Secondly, repeated blood sampling at different ages in ALSPAC, including birth, allowed age-specific cross-sectional analyses. Thirdly, these studies have collected extensive information from obstetric records and reported socio-economic factors allowing adjustment for potential confounders, including birth weight as well as maternal smoking during pregnancy and maternal education. Finally, we used a Mendelian randomization approach as an alternative method to control for unmeasured confounding and examine the direction of observed associations. This provided us with some evidence of a causal relationship between seizures and *BDNF*-linked DNA methylation and pointed towards potential negative consequences of seizures on other health outcomes.

Our results are partially in line with reports showing an involvement of neurotrophins, including *BDNF*, in seizures and epilepsy. Studies conducted in animal models of epilepsy (reviewed in ^42^) observed an upregulation of *BDNF* immediately after experimentally-induced seizures. A study conducted on hippocampal tissue from 40 adult patients affected by mesial-temporal lobe epilepsy showed increased or decreased *BDNF* expression, compared to healthy individuals, depending on the region investigated and on the presence of psychiatric comorbidities ^43^. Similarly, four isoforms of *BDNF* were found to be highly expressed in brain hippocampal tissue from adult epileptic patients compared to healthy controls, although the effect was not explained by changes in DNA methylation measured in the promoters of isoforms IV and VI ^37^. It is to be noted that the association reported in the current study was located further upstream within the first intron of isoforms I, II and III, based on the latest gene characterization ^37, 44^. Moreover, a recent family study investigating genome-wide DNA methylation in peripheral blood, based on 15 trios of parents and their offspring, where the child and one parent, but not the other, were affected by generalized genetic epilepsy, found evidence of neurotrophins involvement, particularly *BDNF*, which was both hyper-and hypomethylated ^45^. In our study, we observed hypermethylation in the *BDNF* gene (in the promoter or within introns, depending on the isoform), which would suggest decreased expression. This is in apparent contrast with some of the previous studies, but in line with our Mendelian randomization analysis that also showed some evidence of seizure-induced hypermethylation in the *BDNF* gene when using generalized genetic epilepsy as the exposure. The apparently contrasting findings might indicate rapid dynamic changes at the molecular level and sub-tissue specificity that need follow-up investigation in biological systems with better time resolution and multi-tissue characterization.

We also discovered associations with other methylation sites that have not been previously observed, specifically in the *MACROD2*, the *PRMT10* genes and two intergenic sites. Single nucleotide polymorphisms within *MACROD2* have previously been associated with autism, although with rather weak evidence ^46^, and other brain-related traits such as intelligence and mathematical abilities ^47^. Individuals with *de novo* mutations in *FBXO11*, an analogous of *PRMT10*, have been reported to show intellectual disability and autism ^48^. Although both genes are predominantly expressed in non-neural tissues, these studies suggest that *MACROD2 and PRMT10* could play a role in brain functioning and their methylation status could plausibly be involved in seizures. However, the involvement of DNA methylation needs to be further confirmed.

The results of this study have to be seen in light of the following limitations. First, despite this being the largest epigenome-wide association study in childhood/adolescence, the sample size was small considering the low prevalence of seizures in the general population. Second, our initial findings did not replicate in data from an independent cohort. However, this could also indicate that the associations are specific to the time window examined in the discovery sample. For instance, the association with *BDNF* methylation was specific to the adolescence time point, while in Generation R data was limited to childhood. Thirdly, we have relied on DNA methylation measured in blood whereas seizures occur within the brain. Availability of brain tissue for epidemiological research is very limited, and future studies could follow-up our findings in animal models or post-mortem brain tissues. Finally, the Mendelian randomization analyses relied on a small number of mQTLs (3 for epilepsy to *BDNF* methylation and 1 for methylation to epilepsy and other health outcomes) and therefore did not allow for sensitivity analyses aimed at ruling out bias due to horizontal pleiotropy.

In conclusion, we show evidence of a link between seizures and DNA methylation in childhood and adolescence, potentially involving *BDNF*. Our study contributes to explaining the relationship between seizures and other health outcomes and suggests that seizures may lead to negative consequences on neurodevelopment and growth via DNA methylation.

## Data Availability

The ALSPAC study website contains details of all the data that is available through a fully searchable data dictionary and variable search tool.

http://www.bris.ac.uk/alspac/researchers/data-access/data-dictionary/

## Acknowledgments

We are extremely grateful to all the families who took part in this study, the midwives for their help in recruiting them, and the whole ALSPAC team, which includes interviewers, computer and laboratory technicians, clerical workers, research scientists, volunteers, managers, receptionists and nurses. The UK Medical Research Council and Wellcome (Grant ref: 102215/2/13/2) and the University of Bristol provide core support for ALSPAC. This publication is the work of the authors and Doretta Caramaschi will serve as guarantors for the contents of this paper. A comprehensive list of grants funding is available on the ALSPAC website (http://www.bristol.ac.uk/alspac/external/documents/grant-acknowledgements.pdf). GWAS data was generated by Sample Logistics and Genotyping Facilities at Wellcome Sanger Institute and LabCorp (Laboratory Corporation of America) using support from 23andMe. Methylation data in the ALSPAC cohort were generated as part of the UK BBSRC funded (BB/I025751/1 and BB/I025263/1) Accessible Resource for Integrated Epigenomic Studies (ARIES, http://www.ariesepigenomics.org.uk). D.C., C.R. and E.W. are funded by UK Medical Research Council (grant numbers: MC_UU_00011/1 and MC_UU_00011/5). C.H. is supported by a 4-year studentship fund from the Wellcome Trust Molecular, Genetic and Lifecourse Epidemiology Ph.D. programme at the University of Bristol (108902/B/15/Z).

The Generation R Study is conducted by the Erasmus Medical Center in close collaboration with the School of Law and Faculty of Social Sciences of the Erasmus University Rotterdam, the Municipal Health Service Rotterdam area, Rotterdam, the Rotterdam Homecare Foundation, Rotterdam and the Stichting Trombosedienst & Artsenlaboratorium Rijnmond (STAR-MDC), Rotterdam. We gratefully acknowledge the contribution of children and parents, general practitioners, hospitals, midwives and pharmacies in Rotterdam. The study protocol was approved by the Medical Ethical Committee of the Erasmus Medical Centre, Rotterdam. Written informed consent was obtained for all participants. The generation and management of the Illumina 450K methylation array data (EWAS data) for the Generation R Study was executed by the Human Genotyping Facility of the Genetic Laboratory of the Department of Internal Medicine, Erasmus MC, the Netherlands. We thank Mr. Michael Verbiest, Ms. Mila Jhamai, Ms. Sarah Higgins, Mr. Marijn Verkerk and Dr. Lisette Stolk for their help in creating the EWAS database. We thank Dr. A.Teumer for his work on the quality control and normalization scripts. The general design of the Generation R Study is made possible by financial support from the Erasmus Medical Center, Rotterdam, the Erasmus University Rotterdam, the Netherlands Organization for Health Research and Development and the Ministry of Health, Welfare and Sport. The EWAS data was funded by a grant from the Netherlands Genomics Initiative (NGI)/Netherlands Organisation for Scientific Research (NWO) Netherlands Consortium for Healthy Aging (NCHA; project nr. 050-060-810), by funds from the Genetic Laboratory of the Department of Internal Medicine, Erasmus MC, and by a grant from the National Institute of Child and Human Development (R01HD068437). J.F.F. has received funding from the European Union’s Horizon 2020 research and innovation programme under grant agreement No 633595 (DynaHEALTH) and from the European Joint Programming Initiative “A Healthy Diet for a Healthy Life” (JPI HDHL, NutriPROGRAM project, ZonMw the Netherlands no.529051022). C.C. has received funding from the European Union’s Horizon 2020 research and innovation programme under the Marie Skłodowska-Curie grant agreement No 707404. This project received funding from the European Union’s Horizon 2020 research and innovation programme (733206, LIFECYCLE).

## Notes

### Competing Interest Statement

The authors have declared no competing interest.

### Author Declarations

All relevant ethical guidelines have been followed and any necessary IRB and/or ethics committee approvals have been obtained.

Any clinical trials involved have been registered with an ICMJE-approved registry such as ClinicalTrials.gov and the trial ID is included in the manuscript.

